# High fibrinogen-prealbumin ratio (FPR) predicts stroke-associated pneumonia

**DOI:** 10.1101/2023.08.09.23293911

**Authors:** Huihua Qiu, Xiaoqian Luan, Enci Mei

**Affiliations:** Department of Neurology, The First Affiliated Hospital of Wenzhou Medical University, Wenzhou 325000, China; Department of Dermatology and Venereology, The First Affiliated Hospital of Wenzhou Medical University, Wenzhou 325000, China

**Author notes:** **Correspondence:** Enci Mei, Department of Dermatology and Venereology, The First Affiliated Hospital of Wenzhou Medical University, Wenzhou, 325000, Zhejiang Province, China.

**Keywords:** pneumonia, ischemia, fibrinogen, prealbumin, stroke

## Abstract

**Background:** Stroke-associated pneumonia (SAP) is a common complication in acute ischemic stroke patients. Although both single markers of fibrinogen and prealbumin are found to be associated with stroke and pneumonia, fibrinogen-prealbumin ratio (FPR) is a novel and comprehensive indicator that has not been explored in acute ischemic stroke patients. Besides, no study has explored the relationship between SAP and FPR. This study aims to explore whether FPR is higher in acute ischemic stroke patients and whether FPR is associated with SAP.

**Methods:** A total of 902 acute ischemic stroke patients participated in this study. Meanwhile, 146 healthy controls were also recruited. Fibrinogen and prealbumin were measured within 24 hours on admission. FPR was calculated after dividing fibrinogen (g/L) by prealbumin (mg/L)× 1000. SAP was defined according to the modified Centers for Disease Control criteria.

**Results:** In this study, 121 patients were diagnosed with SAP. FPR was significantly higher in both non-SAP patients and SAP patients than in healthy controls. In binary logistic regression analysis, we found that FPR was significantly higher in SAP group than non-SAP group (15.97[11.72-24.34] vs. 11.81[9.27-15.64]; P < 0.001) after adjusting for confounders. Besides, FPR (>18.22) was independently associated with SAP (OR3.028; 95% CI:1.607-5.706; P = 0.001). Moreover, diabetes mellitus, NIHSS score, dysphagia, leukocyte count and hs-CRP were independently correlated with SAP.

**Conclusion:** Higher FPR was observed in acute ischemic stroke patients compared to healthy controls and high FPR significantly increased the risk of SAP. Patients with high FPR should be paid more attention by physicians.

## Introduction

Stroke-associated pneumonia (SAP) is a common complication in acute ischemic stroke patients, approximately 14% of patients develop SAP after stroke ^1, 2^. SAP is shown to be associated with poor functional prognosis, poor quality of life and early mortality after stroke^3^. Therefore, it is of great significance for seeking the early identification and treatment of SAP. However, the underlying mechanisms of SAP remain unclear.

Previous research had shown that acute ischemic stroke would trigger a local inflammatory immune response, which led to immunosuppression and an increased risk of stroke-associated infection ^4, 5^. Numerous factors had been discovered to be associated with SAP. Dysphagia, impaired mobility and immunodepression were identified as risk factors of SAP ^4^. Additionally, several inflammatory biomarkers were considered to be predictors of SAP, such as fibrinogen, neutrophil-to-lymphocyte ratio (NLR), monocyte-lymphocyte ratio (MLR) ^6, 7^.

As a plasma glycoprotein, fibrinogen is synthesized in the liver. It is composed of three pairs of non-identical polypeptide chains including the α-, β-, and γ-chains ^8^. Fibrinogen modulates the thrombosis and coagulation process, which is involved in cell attachment, hemostasis, as well as systemic inflammatory reactions ^9^. Clinicians have recognized associations of fibrinogen with acute ischemic stroke and pneumonia ^7, 10^. Prealbumin is a protein produced in the liver and choroid plexus, playing a role in the transport of thyroid hormone as well as vitamin A ^11^. During inflammation and malnutrition which are commonly seen in acute ischemic stroke, the synthesis rate of prealbumin would decline due to reprioritization of the synthesis to acute-phase proteins such as C-reactive protein (CRP) ^12^. Moreover, previous studies have found that prealbumin is independently associated with acute ischemic stroke and pneumonia ^13–15^.

As a novel and effective biomarker, fibrinogen-prealbumin ratio (FPR) could balance the effects of inflammation and nutrition, which reflects biological status of patients more comprehensively compared to single markers of fibrinogen and prealbumin ^16^. However, to date, FPR has not been explored in acute ischemic stroke patients and no study has been conducted to explore the relationship between SAP and FPR. Therefore, in this study, whether FPR was higher in patients with acute ischemic stroke and whether FPR was associated with SAP were evaluated.

## Materials and Methods

### Study participants

Patients were selected from the First Affiliated Hospital of Wenzhou Medical University from the year of 2016 to 2021. Inclusion criteria included: 1) 18-80 years old; 2) acute ischemic stroke within 7 days and confirmed by computed tomography (CT) or magnetic resonance imaging (MRI). The exclusion criteria were as follows: 1) transient ischemic attack 2) active infection within two weeks before admission, 3) severe liver or kidney disease, 4) severe neurological diseases, such as epilepsy, Parkinson’s disease and hydrocephalus, 5) history of malignancy, 6) lack of complete laboratory data. At last, 902 patients participated in our study (Fig. 1). Meanwhile, there were 146 healthy controls recruited from our hospital during physical check-ups, who were confirmed without severe physical diseases, including stroke and pneumonia. This study was approved by the Medical Ethics Committee of the First Affiliated Hospital of Wenzhou Medical University. Each patient had signed the informed consent form. The screening of the flow chart is shown in Figure 1.

**Fig. 1.**
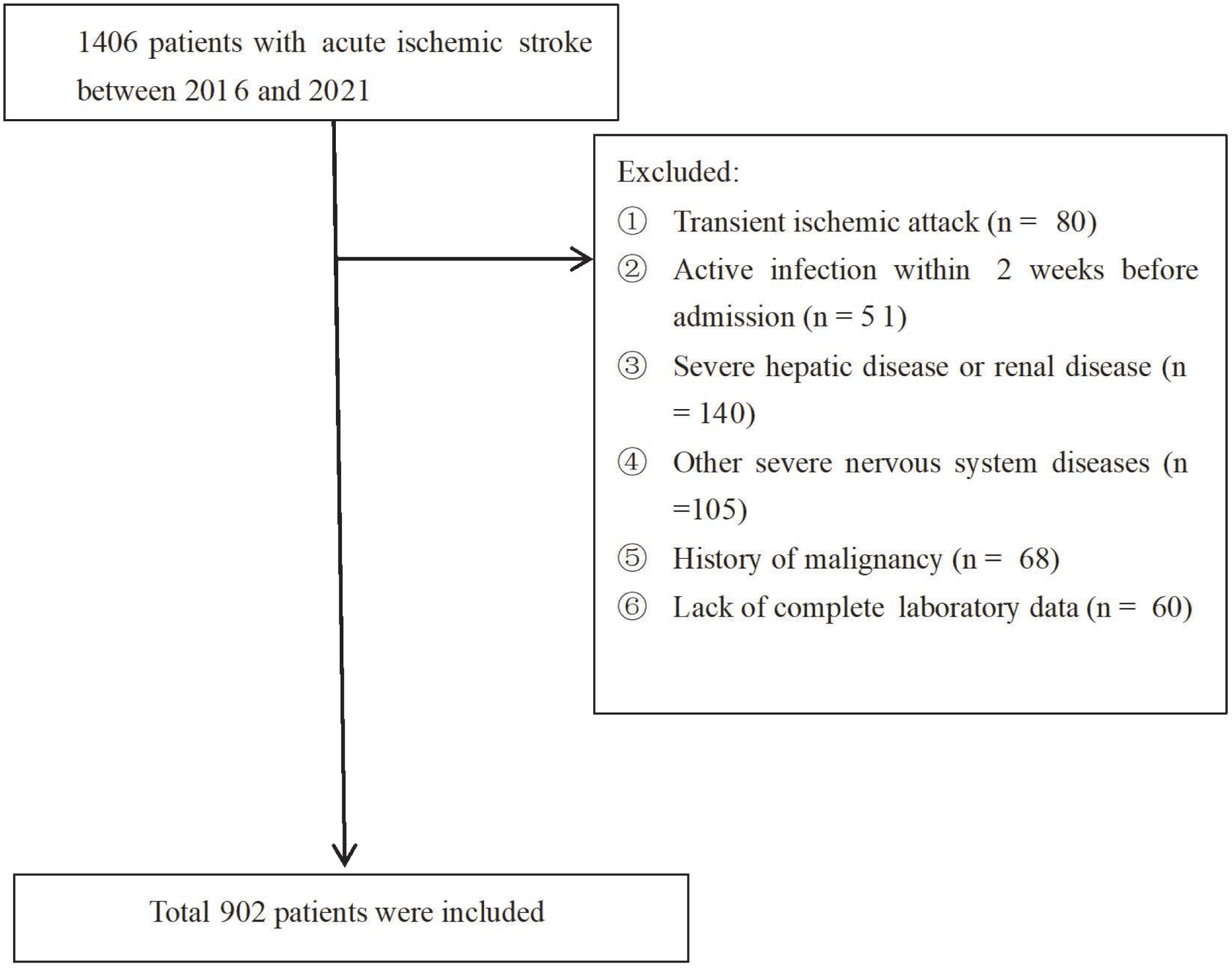
Study flow diagram.

### Clinical variables

Pneumonia before acute ischemic stroke and community-acquired pneumonia were excluded from this study. According to the modified Centers for Disease Control and Prevention criteria for hospital-acquired Pneumonia ^17^, lower respiratory tract infection within the first 7 days after stroke onset was diagnosed as SAP. To reduce the burden of CT examinations or X-rays, both the definite and probable SAP were considered as SAP ^18^. SAP was assessed by two experienced neurologists who were blinded to the results.

Demographic data were collected based on the patients’ medical history, such as sex, age, diabetes mellitus, hypertension, hyperlipidemia, coronary heart disease, stroke history. Besides, stroke subtypes were classified according to TOAST criteria ^19^. Clinical factors were recorded on admission, including dysphagia, baseline NIHSS score and use of thrombolysis. Stroke severity was assessed by trained neurologists using the National Institutes of Health Stroke score (NIHSS) score on admission ^20^. Dysphagia was confirmed by rehabilitative therapist in our hospital within the first day of admission via a 30ml water swallowing test (WST) ^21^. When the patient sat upright, the therapist gave the patient 5ml of water twice, and then they asked the patient to drink the rest of the water. Any cough or voice change after drinking was considered as dysphagia. In addition, A2DS2 score was an effective predictor of SAP, ranging from 0 to 10 points. A2DS2 score consists of atrial fibrillation (1 points), gender (male 1 points), age (75 years 1 points), dysphagia (2 points) and stroke severity at admission (NIHSS score 0–4:0 point; NIHSS score 5–15:3 points; NIHSS score ≥16:5 points)^22^. Blood samples were collected within 24 hours after admission.

### Laboratory examination

The laboratory examination was performed on admission. Serum prealbumin was measured by an immunoturbidimetric method on immunoassay analyzer (Beckman Coulter Inc., Fullerton, CA, USA) with the same batch of reagents. The NLR and FPR were defined as neutrophils (10^9^/L)/lymphocytes (10^9^/L) and fibrinogen (g/L) by prealbumin (mg/L)× 1000 ^16^.

### Statistical analysis

Continuous variables were expressed as mean ± standard deviation (SD) or median (quartile range, IQR) according to normal distribution. Besides, categorical variables were demonstrated in the form of counts and percentages. Chi-squared test was used to compare the categorical variables between the SAP group and the non-SAP group while Student’s t test and Mann–Whitney test were used to compare the continuous variables between the two groups. Pearson correlation coefficients were applied to identify the variables which were correlated with FPR. In order to find out the variables which were correlated with SAP, binary logistic regression analysis was conducted, in which variables with *P<*0.05 in the univariate analysis between the SAP group and the non-SAP group were included. Moreover, an additional sensitivity analysis was conducted by using A2DS2 score as a confounding variable instead of age, atrial fibrillation, dysphagia and NIHSS score to confirm the main results of this study ^22, 23^. To explore the predictive power of FPR, leukocyte count, NLR, hs-CRP, albumin and A2DS2 score for SAP, the receiver-operating characteristic (ROC) was applied in this study. Furthermore, A2DS2 score was used to evaluate the additive effect of FPR on predicting SAP, according to the method used in previous study ^24^. IBM SPSS Statistics 19.0 was used for statistical analysis and two-tailed P-values < 0.05 were considered to be statistically significant.

## Results

### Baseline characteristics

Clinical and demographic characteristics of the samples under this study are shown in Table 1. In the study population, 121 patients (13.41%) were diagnosed with SAP. There was no significant intergroup difference in terms of sex and BMI among SAP group, non-SAP group and healthy controls. The healthy controls had no significant difference in age compared with SAP group and non-SAP group. Furthermore, FPR was significantly higher in non-SAP patients and SAP patients than in healthy controls while prealbumin was significantly lower in non-SAP patients and SAP patients than in healthy controls. Besides, FIB in SAP group was higher than that in non-SAP group and healthy controls.

**Table 1.**
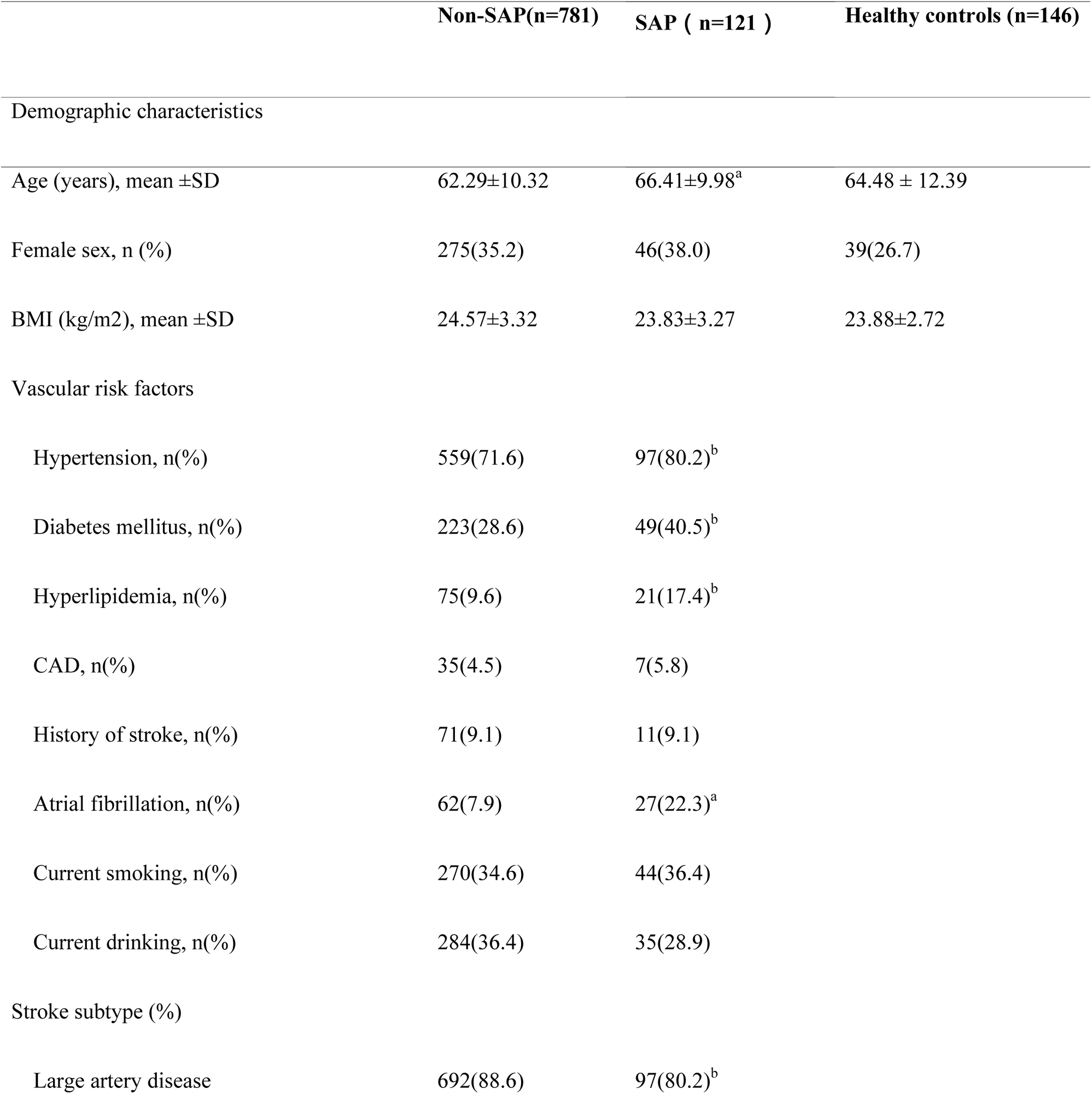
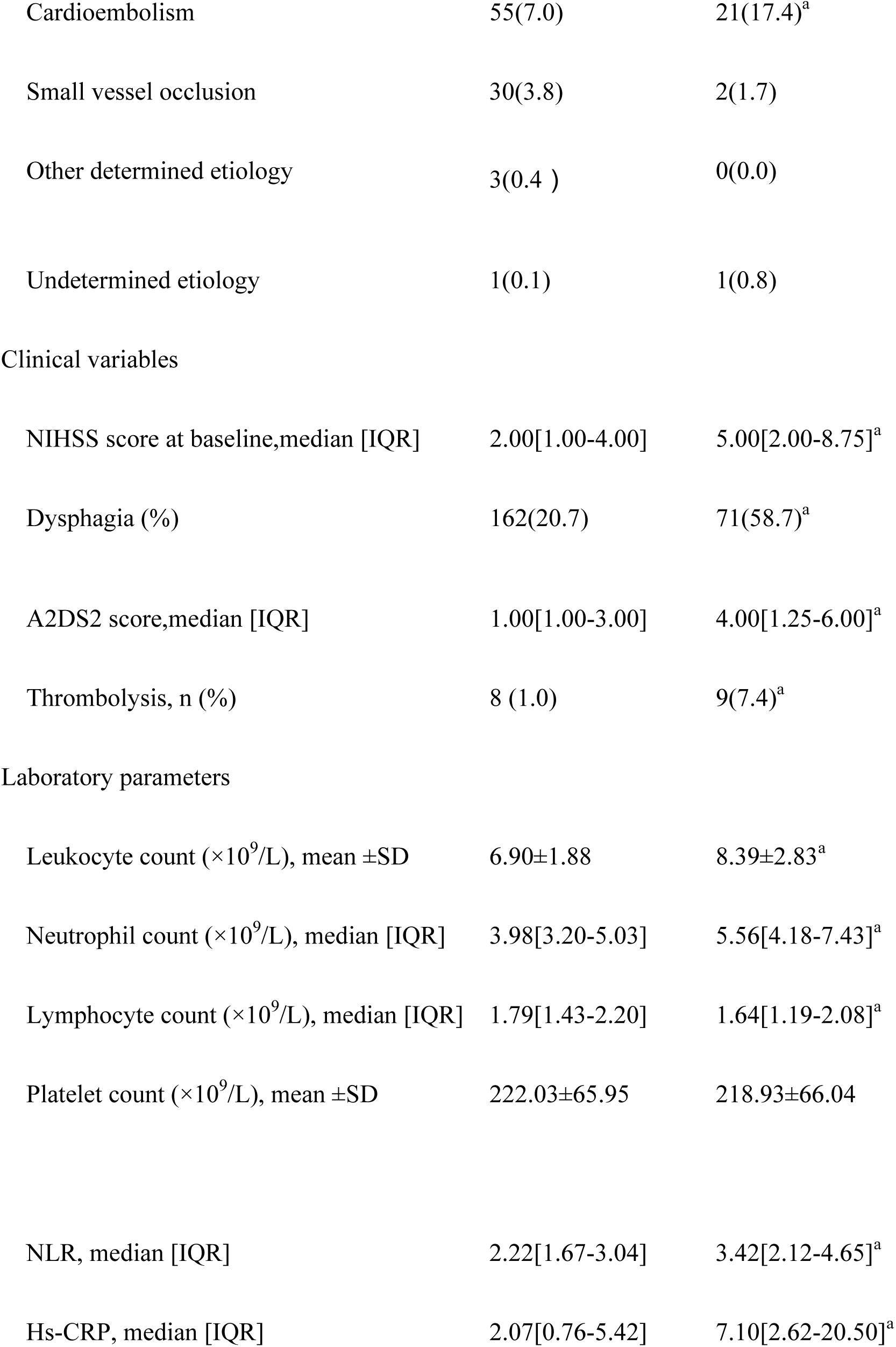
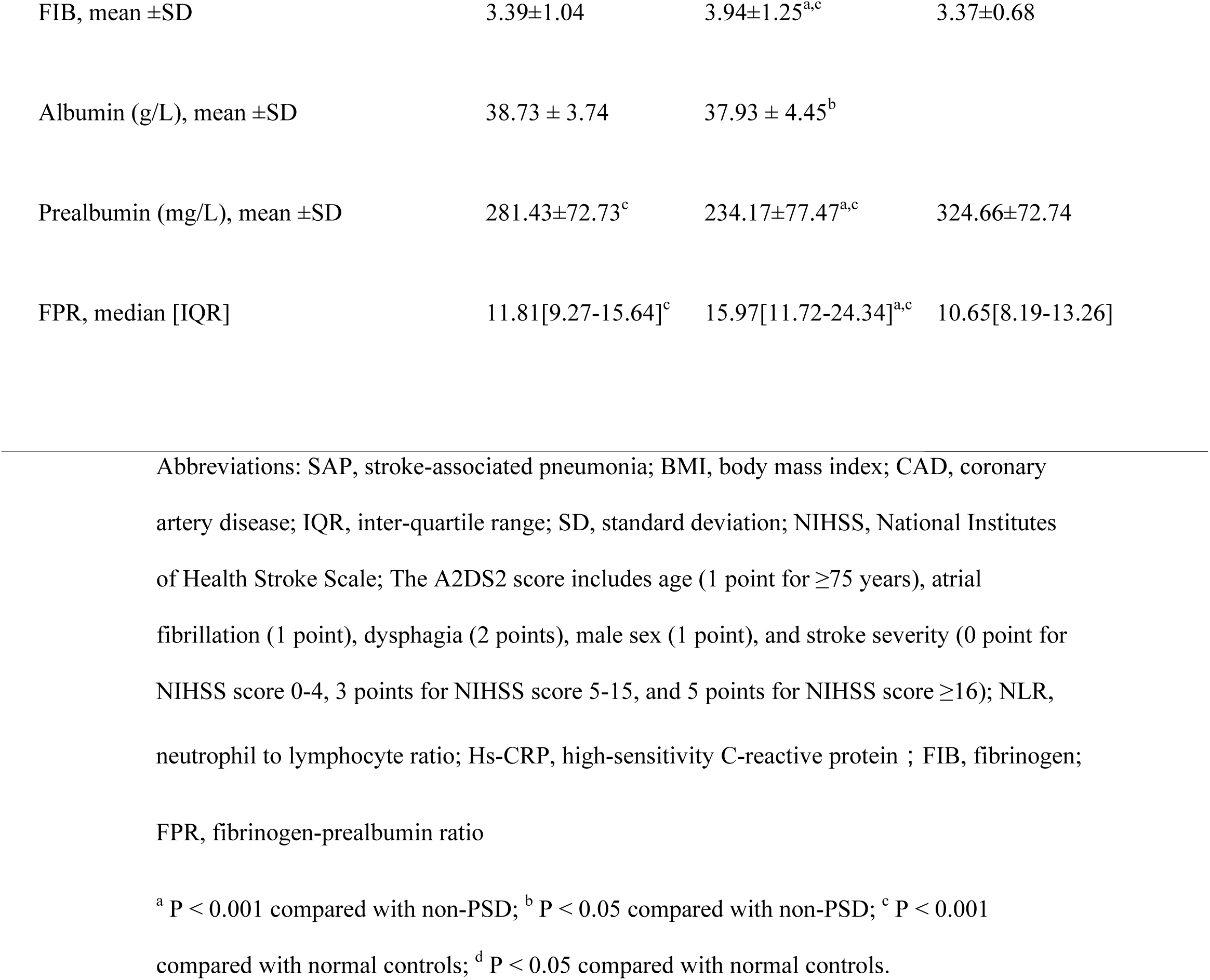
Clinical and demographic characteristics of the samples under study.

|  | Non-SAP(n=781) | SAP ( n=121 ) | Healthy controls (n=146) |
| --- | --- | --- | --- |
| Demographic characteristics |  |  |  |
| Age (years), mean ±SD | 62.29±10.32 | 66.41±9.98<sup>a</sup> | 64.48 ± 12.39 |
| Female sex, n (%) | 275(35.2) | 46(38.0) | 39(26.7) |
| BMI (kg/m<sup>2</sup>), mean ±SD | 24.57±3.32 | 23.83±3.27 | 23.88±2.72 |
| Vascular risk factors |  |  |  |
| Hypertension, n(%) | 559(71.6) | 97(80.2)<sup>b</sup> |  |
| Diabetes mellitus, n(%) | 223(28.6) | 49(40.5)<sup>b</sup> |  |
| Hyperlipidemia, n(%) | 75(9.6) | 21(17.4)<sup>b</sup> |  |
| CAD, n(%) | 35(4.5) | 7(5.8) |  |
| History of stroke, n(%) | 71(9.1) | 11(9.1) |  |
| Atrial fibrillation, n(%) | 62(7.9) | 27(22.3)<sup>a</sup> |  |
| Current smoking, n(%) | 270(34.6) | 44(36.4) |  |
| Current drinking, n(%) | 284(36.4) | 35(28.9) |  |
| Stroke subtype (%) |  |  |  |
| Large artery disease | 692(88.6) | 97(80.2)<sup>b</sup> |  |

|  |  |  |
| --- | --- | --- |
| Cardioembolism | 55(7.0) | 21(17.4)<sup>a</sup> |
| Small vessel occlusion | 30(3.8) | 2(1.7) |
| Other determined etiology | 3(0.4 ) | 0(0.0) |
| Undetermined etiology | 1(0.1) | 1(0.8) |
| Clinical variables |  |  |
| NIHSS score at baseline,median [IQR] | 2.00[1.00-4.00] | 5.00[2.00-8.75]<sup>a</sup> |
| Dysphagia (%) | 162(20.7) | 71(58.7)<sup>a</sup> |
| A2DS2 score,median [IQR] | 1.00[1.00-3.00] | 4.00[1.25-6.00]<sup>a</sup> |
| Thrombolysis, n (%) | 8 (1.0) | 9(7.4)<sup>a</sup> |
| Laboratory parameters |  |  |
| Leukocyte count ($\times 10^9/L$), mean ±SD | 6.90±1.88 | 8.39±2.83<sup>a</sup> |
| Neutrophil count ($\times 10^9/L$), median [IQR] | 3.98[3.20-5.03] | 5.56[4.18-7.43]<sup>a</sup> |
| Lymphocyte count ($\times 10^9/L$), median [IQR] | 1.79[1.43-2.20] | 1.64[1.19-2.08]<sup>a</sup> |
| Platelet count ($\times 10^9/L$), mean ±SD | 222.03±65.95 | 218.93±66.04 |
| NLR, median [IQR] | 2.22[1.67-3.04] | 3.42[2.12-4.65]<sup>a</sup> |
| Hs-CRP, median [IQR] | 2.07[0.76-5.42] | 7.10[2.62-20.50]<sup>a</sup> |
| FIB, mean ±SD | 3.39±1.04 | 3.94±1.25<sup>a,c</sup> | 3.37±0.68 |
| Albumin (g/L), mean ±SD | 38.73 ± 3.74 | 37.93 ± 4.45<sup>b</sup> | |
| Prealbumin (mg/L), mean ±SD | 281.43±72.73<sup>c</sup> | 234.17±77.47<sup>a,c</sup> | 324.66±72.74 |
| FPR, median [IQR] | 11.81[9.27-15.64]<sup>c</sup> | 15.97[11.72-24.34]<sup>a,c</sup> | 10.65[8.19-13.26] |

Abbreviations: SAP, stroke-associated pneumonia; BMI, body mass index; CAD, coronary artery disease; IQR, inter-quartile range; SD, standard deviation; NIHSS, National Institutes of Health Stroke Scale; The A2DS2 score includes age (1 point for $\geq 75$ years), atrial fibrillation (1 point), dysphagia (2 points), male sex (1 point), and stroke severity (0 point for NIHSS score 0-4, 3 points for NIHSS score 5-15, and 5 points for NIHSS score $\geq 16$ ); NLR, neutrophil to lymphocyte ratio; Hs-CRP, high-sensitivity C-reactive protein ; FIB, fibrinogen; FPR, fibrinogen-prealbumin ratio
<sup>a</sup> P < 0.001 compared with non-PSD; <sup>b</sup> P < 0.05 compared with non-PSD; <sup>c</sup> P < 0.001 compared with normal controls; <sup>d</sup> P < 0.05 compared with normal controls.

The level of FPR was significantly higher in SAP group than that in non-SAP group (15.97[11.72-24.34] vs. 11.81[9.27-15.64]; P < 0.001). Compared with the non-SAP group, the SAP group was elder and had a higher proportion of hypertension, diabetes, hyperlipidemia, atrial fibrillation, cardiogenic thrombus, dysphagia and thrombolysis. Moreover, NIHSS score, A2DS2 score, leukocyte count, neutrophil count, NLR and hs-CRP levels in SAP group were higher than those in non-SAP group. In addition, the proportion of large artery disease, lymphocyte count, albumin and prealbumin level in SAP group were lower than those in non-SAP group (Table 1).

In correlation analysis, we found that FPR was positively correlated with age, hs-CRP, leukocyte count, platelet count, NLR, A2DS2 score and NIHSS score (all p<0.05), while FPR was negatively correlated with lymphocyte count and albumin (p<0.05) (Table 2).

**Table 2.**
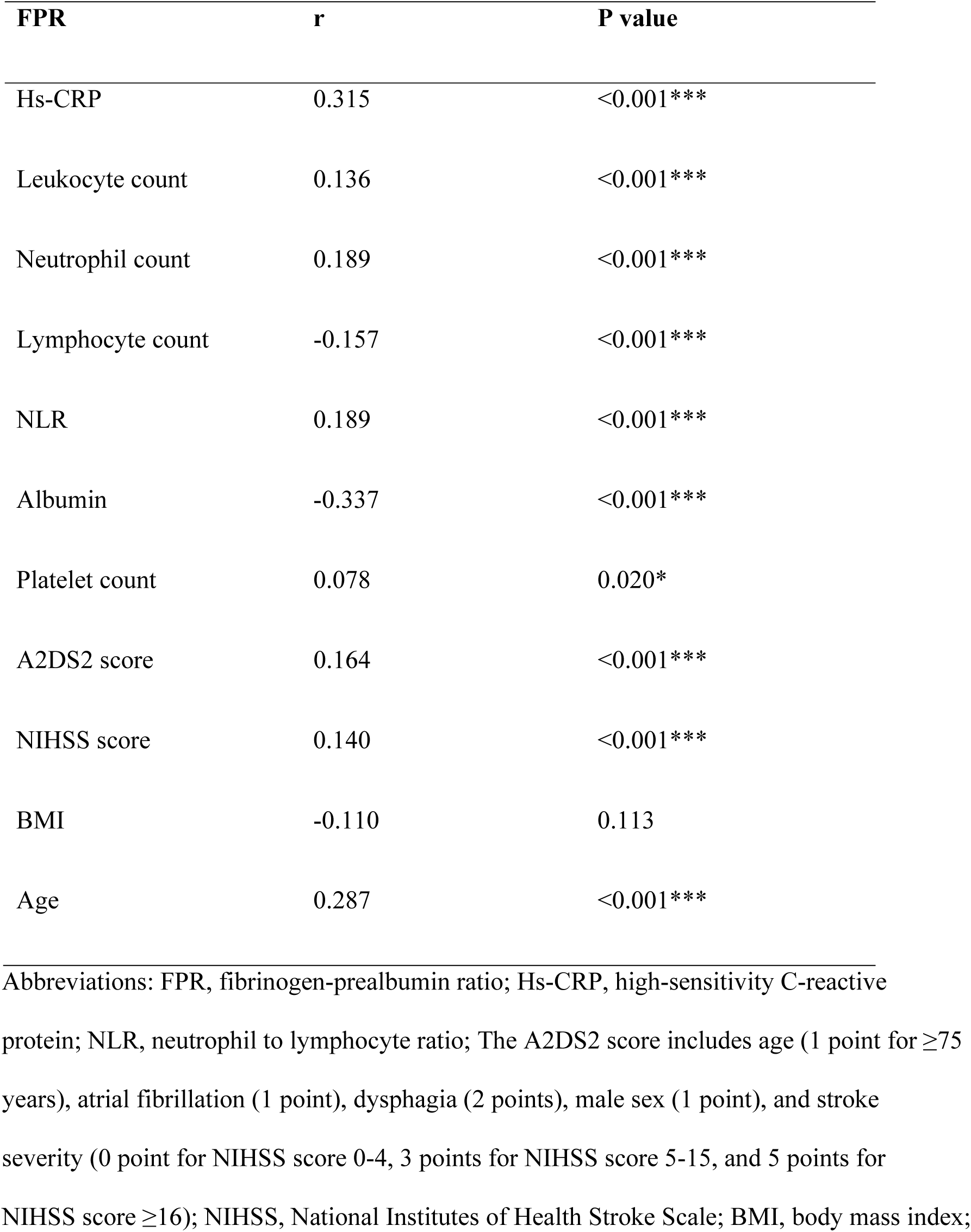
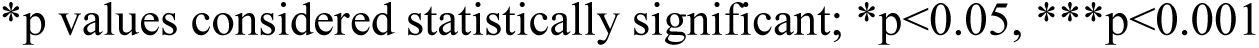
The evaluation of the correlation coefficient between FPR and other potential confounding variables.

| **FPR** | **r** | **P value** |
| --- | --- | --- |
| Hs-CRP | 0.315 | <0.001*** |
| Leukocyte count | 0.136 | <0.001*** |
| Neutrophil count | 0.189 | <0.001*** |
| Lymphocyte count | -0.157 | <0.001*** |
| NLR | 0.189 | <0.001*** |
| Albumin | -0.337 | <0.001*** |
| Platelet count | 0.078 | 0.020* |
| A2DS2 score | 0.164 | <0.001*** |
| NIHSS score | 0.140 | <0.001*** |
| BMI | -0.110 | 0.113 |
| Age | 0.287 | <0.001*** |

Abbreviations: FPR, fibrinogen-prealbumin ratio; Hs-CRP, high-sensitivity C-reactive protein; NLR, neutrophil to lymphocyte ratio; The A2DS2 score includes age (1 point for $\geq 75$ years), atrial fibrillation (1 point), dysphagia (2 points), male sex (1 point), and stroke severity (0 point for NIHSS score 0-4, 3 points for NIHSS score 5-15, and 5 points for NIHSS score $\geq 16$ ); NIHSS, National Institutes of Health Stroke Scale; BMI, body mass index;
\*p values considered statistically significant; \*p<0.05, \*\*\*p<0.001

### The relationship between FPR and SAP

As demonstrated in Table 3, the level of FPR on admission was independently correlated with SAP (OR 1.091; 95% CI: 1.045-1.139; P<0.001) in binary logistic regression analysis. Moreover, FPR remained independently correlated with SAP after replacing initial NIHSS score, age, atrial fibrillation and dysphagia with A2DS2 score as a sensitivity analysis (OR 1.095; 95% CI (1.050-1.142); P<0.001). In addition, dysphagia (OR 2.301; 95% CI: 1.230-4.306; P =0.009), diabetes mellitus (OR1.992; 95% CI: 1.075-3.694; P =0.029), NIHSS score (OR1.124; 95% CI: 1.032-1.224; P =0.007), leukocyte count (OR1.194; 95% CI: 1.047-1.360; P =0.008) and hs-CRP value (OR1.015; 95% CI: 1.005-1.026; P =0.005) were also independently related to SAP.

**Table 3.** Binary logistic model of the clinical determinants of SAP.

|  | Model 1 † (n=902) |  | Model 2 ‡ (n=902) |  |
| --- | --- | --- | --- | --- |
|  | Adjusted OR (95%CI) | P Value | Adjusted OR (95%CI) | P Value |
| Age |  | 0.295 | ... | ... |
| Hypertension |  | 0.526 |  | 0.711 |
| Diabetes mellitus | 1.992(1.075-3.694) | 0.029* | 2.050(1.117-3.764) | 0.021* |
| Hyperlipidemia |  | 0.079 | 2.333(1.087-5.009) | 0.030* |
| Atrial fibrillation |  | 0.067 | ... | ... |
| Stroke subtype |  | 0.273 |  | 0.478 |
| NIHSS score at baseline | 1.124(1.032-1.224) | 0.007* | ... | ... |
| Dysphagia | 2.301(1.230-4.306) | 0.009* | ... | ... |
| A2DS2 score | ... | ... | 1.439(1.250-1.656) | <0.001*** |
| Thrombolysis |  | 0.674 |  | 0.468 |
| Leukocyte count | 1.194(1.047-1.360) | 0.008* | 1.165(1.021-1.330) | 0.023* |
| NLR |  | 0.070 |  | 0.070 |
| Hs-CRP | 1.015(1.005-1.026) | 0.005* | 1.014(1.004-1.025) | 0.007* |
| Albumin |  | 0.659 |  | 0.698 |
| FPR | 1.091(1.045-1.139) | <0.001*** | 1.095(1.050-1.142) | <0.001*** |

Abbreviations: SAP, stroke-associated pneumonia; OR: odds ratio; CI: confidence interval; NIHSS, National Institutes of Health Stroke Scale; The A2DS2 score includes age (1 point for $\geq 75$ years), atrial fibrillation (1 point), dysphagia (2 points), male sex (1 point), and stroke severity (0 point for NIHSS score 0-4, 3 points for NIHSS score 5-15, and 5 points for NIHSS score $\geq 16$ ); NLR, neutrophil to lymphocyte ratio; Hs-CRP, high-sensitivity C-reactive protein; FPR, fibrinogen-prealbumin ratio
†Adjusted with $P < 0.05$ in univariate analysis
‡Adjusted with $P < 0.05$ in the univariate analysis and A2DS2
\*p values considered statistically significant; \* $p < 0.05$ , \*\*\* $p < 0.001$

As shown in Figure 2, after using the ROC curve, we found that the FPR at admission of 18.22 could predict the development of SAP, with an area under the curve (AUC) of 0.696 (95% CI: 0.665-0.726), with a sensitivity of 47.11 % and a specificity of 83.87 %. At the same time, we found that the predictive power of hs-C reactive protein (0.747 [0.715-0.777]) was higher than that of FPR (0.696 [0.665-0.726]), leukocyte count (0.675 [0.641-0.707]), NLR (0.711[0.678-0.742]), albumin (0.548 [0.513-0.583]) and A2DS2 (0.728 [0.696-0.759]). In addition, the predictive power of FPR, hs-CRP levels, NLR, leukocyte count and A2DS2 were all higher than that of albumin (all P < 0.05). However, there was no significant difference among FPR, hs-CRP levels, NLR, leukocyte count and A2DS2 (all P > 0.05).

**Fig. 2.**
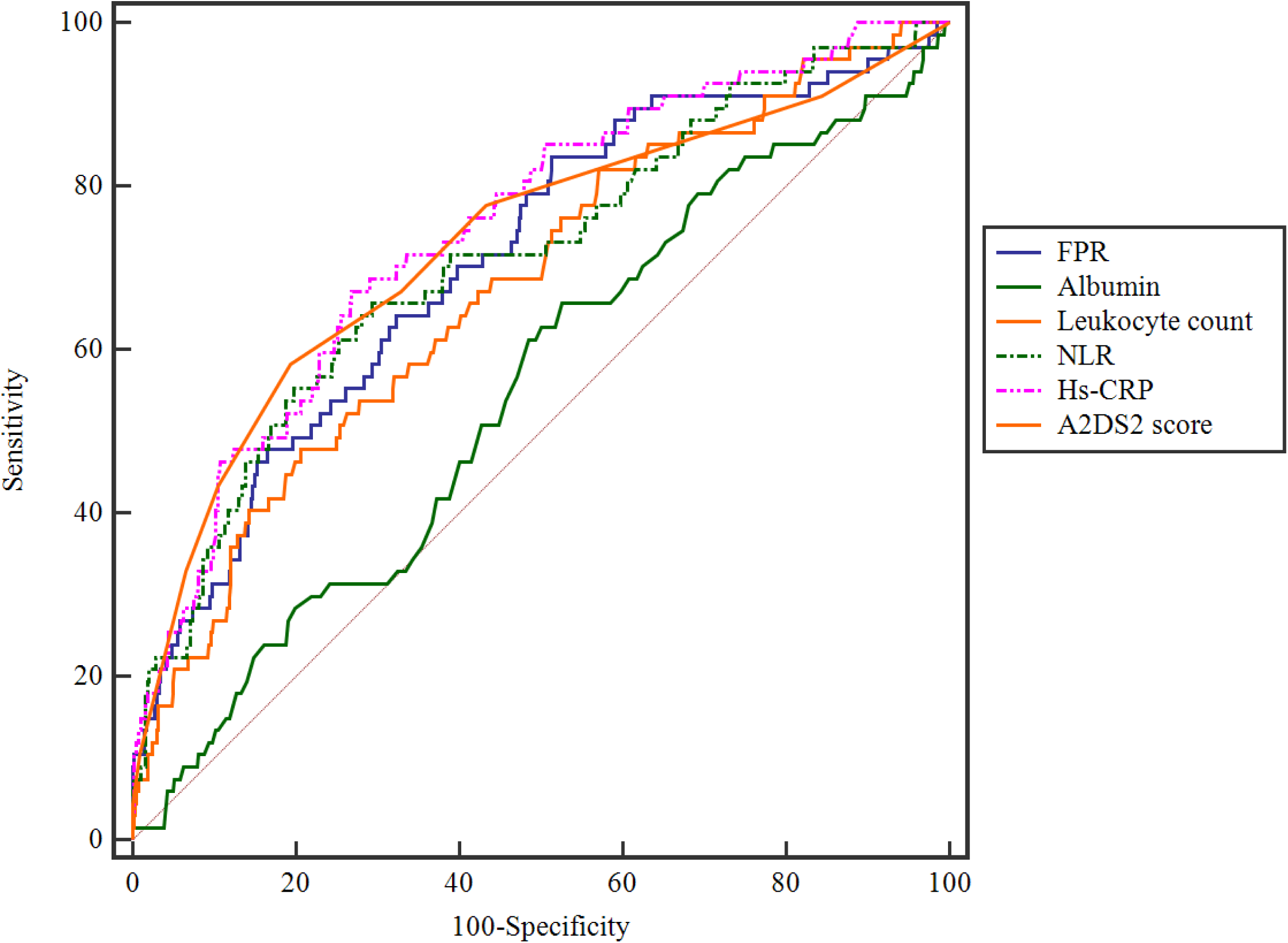
Comparison of predictive power between FPR, A2DS2 score and other biomarkers in the prediction of SAP.

Moreover, as shown in Figure 3, the area under A2DS2 score curve increased significantly while combining FPR with A2DS2 score (0.821[0.794-0.846]; P<0.001). At the same time, the area under the curve of FPR combined with A2DS2 score was higher than that of leukocyte count combined with A2DS2 score (0.766 [0.735-0.794]; P > 0.05), NLR combined with A2DS2 score (0.762 [0.731-0.791]; P > 0.05), hs-CRP combined with A2DS2 score (0.793 [0.764-0.821]; P > 0.05) and albumin combined with A2DS2 score (0.733 [0.701-0.763]; P = 0.002). In addition, while replacing FPR with FPR >18.22 in the binary logistic model, there was an increased risk of SAP (OR3.028; 95% CI:1.607-5.706; P = 0.001) after adjustment for confounding factors (Supplemental Table I).

**Fig. 3.**
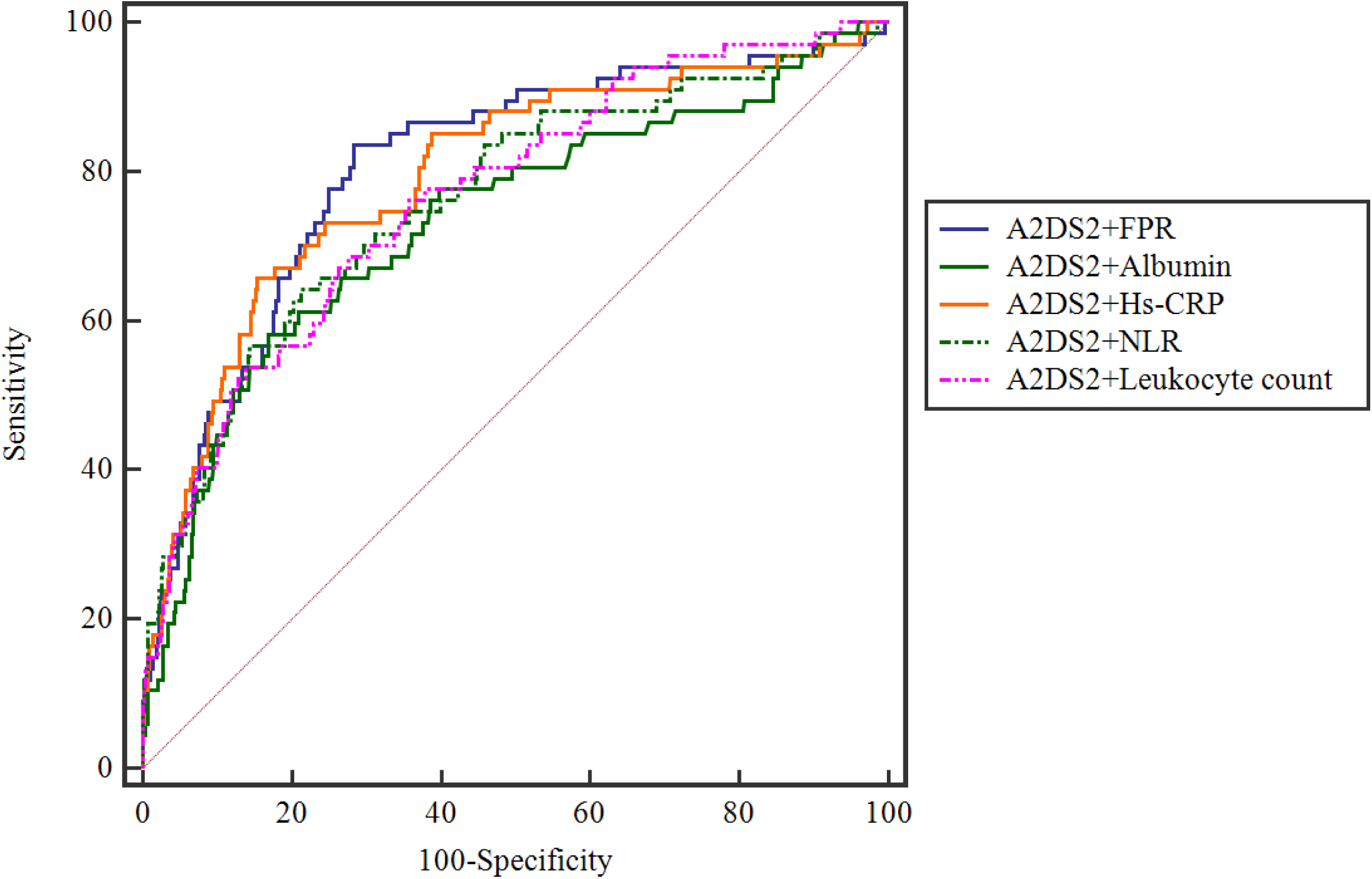
Comparison of predictive power between FPR and other biomarkers combining with A2DS2 score in the prediction of SAP.

## Discussion

To the best of our knowledge, our study is the first to compare the difference of FPR level between acute ischemic stroke patients and healthy controls. Moreover, this study is the first to explore the relationship between FPR and SAP. Our results showed that FPR was higher in acute ischemic stroke patients compared to healthy controls and high level of FPR was independently associated with SAP.

Previous researches had demonstrated that high fibrinogen level was associated with an increased risk of acute ischemic stroke ^25, 26^. Meanwhile, prealbumin level is an important biomarker reflecting the nutritional status of patients ^27, 28^, which is showed to be strongly correlated with acute ischemic stroke ^29^. In this study, increased FPR was observed in acute ischemic stroke patients compared to healthy controls, which was consistent with previous studies.

Our results demonstrated that FPR was independently correlated with SAP. Furthermore, patients with high level of FPR (>18.22) had a 3-fold increase in the risk of developing SAP (OR3.028; 95% CI:1.607-5.706; P = 0.001). Besides, in this study, FPR was found to be positively correlated with hs-CRP, leukocyte count and NLR. Meanwhile, FPR was negatively correlated with albumin. These findings demonstrated that FPR could indicate the level of inflammation as well as the nutritional status of patients. Thus, FPR reflects patients’ biological status more comprehensively than single markers of fibrinogen and prealbumin. Moreover, while combined with A2DS2, FPR could improve the diagnostic efficiency of SAP, which was higher than inflammatory indicators (leukocyte count, NLR and hs-CRP) as well as nutritional indicator (albumin). These findings were consistent with previous research^16^.

FPR may affect SAP through different biological mechanisms. Firstly, there is an inflammatory response after acute ischemic stroke, which is strongly associated with the susceptibility to infections including SAP ^30, 31^. High fibrinogen level was showed to be correlated with inflammatory response in acute ischemic stroke patients ^25^. Meanwhile, prealbumin level could reflect minor changes caused by malnutrition in a short period as a result of its short half-life ^32^. Therefore, prealbumin level on admission might indicate the nutritional status of patients ^27, 28^. Several studies have found that malnutrition has negative effect on inflammatory response ^33, 34^, which is strongly associated with pneumonia ^35, 36^. Interestingly, higher fibrinogen level and lower prealbumin level were found in SAP group compared to non-SAP group in our study. Thus, FPR might contribute to the development of SAP via inflammatory response. Secondly, stroke severity could be a connector between FPR and SAP. Previous studies had demonstrated that high fibrinogen level was independently correlated with severe stroke ^37, 38^. Meanwhile, low prealbumin level was also found to have a close relationship with stroke severity ^39^. Furthermore, stroke severity was shown to contribute to the development of SAP ^40^. In this study, FPR was positively correlated with NIHSS score, which was consistent with previous research ^37, 39^. Therefore, FPR may act as a marker of stroke severity in patients who are vulnerable to SAP.

Thirdly, patients with high FPR level might have reduced nutritional levels and a reduced burden of inflammation, which was correlated with immunosuppression ^41^. Previous research found immunosuppression played an important role in the development of infectious complications after acute ischemic stroke ^42^. Lymphocyte count was reported to be a useful indicator of immunosuppression ^43, 44^. Interestingly, in this study, lymphocyte count was significantly lower in SAP patients, which indicated that immunosuppression was involved in SAP. Moreover, there was a negative correlation between FPR and lymphocyte count. Therefore, immunosuppression might participate in the development of SAP among patients with high FPR.

This study has several limitations. Firstly, FPR was measured only once on admission. Further research is needed to explore the dynamic relationship between FPR and SAP. Secondly, this study was single-center, which might limit conclusions. In future, multicenter clinical research is essential to validate the relationship between SAP and FPR. Thirdly, oral status and other nutritional markers were lacked in our study. Finally, both definite and probable SAP were included as a single variable. Since probable SAP had no radiological evidence, several differential diagnoses might mimic SAP, including viral infection and inflammation. However, the main results of our study remained consistent when we defined SAP as only definite SAP (Supplemental Table II-III). Although patients with probable SAP accounted for only 17.4% of SAP cases in this study, the possibility of overestimation of SAP prevalence should be considered.

In conclusion, this study showed that FPR in acute ischemic stroke patients was higher than that in healthy controls. Moreover, high level of FPR on admission was independently associated with SAP. In conjunction with clinical manifestation, patients with high FPR level were at high risk of SAP and should be paid more attention by physicians. Moreover, our study demonstrated that FPR could not only reflect the level of inflammation in patients, but also indicate the nutritional status of patients. Thus, both anti-inflammation and nutritional supplements might be important to for patients with high FPR level to prevent SAP.

## Data Availability

The data of this study are available on request from the corresponding author. The data are not publicly available due to privacy or ethical restrictions.

## Statements

## Acknowledgement

We are greatly indebted to the staff as well as the patients for their contributions during this work.

## Ethical standards

Our study was approved by the medical ethics committee of the First Affiliated Hospital of Wenzhou Medical University and had abided by the ethical guidelines of the 1964 Declaration of Helsinki and its later amendments. Each patient gave the signature of inform consent form.

## Conflicts of interest

The authors declared that they have no conflicts of interest to this work.

## Funding Sources

This work was supported by Wenzhou Municipal Sci-Tech Bureau Program [grant number Y2020421].

## Author’s contributions

Authors Enci Mei, Huihua Qiu designed the study and wrote the protocol. Author Huihua Qiu and Xiaoqian Luan conducted literature searches and provided summaries of previous studies. Author Huihua Qiu conducted the statistical analysis and wrote the first draft of the manuscript. All authors contributed to and have approved the final manuscript.

## Availability of data

### Code availability

The processed data required to reproduce these results could not be shared at this time as the data also forms part of an ongoing study.

---

**Supplemental Table I.** Binary logistic model of the clinical determinants of SAP, replacing FPR with FPR 18.22

|  | Model 1 † (n=902) |  | Model 2 ‡ (n=902) |  |
| --- | --- | --- | --- | --- |
|  | Adjusted OR | P Value | Adjusted OR | P Value |
| Age |  | 0.295 | ... | ... |
| Hypertension |  | 0.526 |  | 0.711 |
| Diabetes mellitus | 1.992(1.075-3.694) | 0.029* | 2.050(1.117-3.764) | 0.021* |
| Hyperlipidemia |  | 0.079 | 2.333(1.087-5.009) | 0.030* |
| Atrial fibrillation |  | 0.067 | ... | ... |
| Stroke subtype |  | 0.273 |  | 0.478 |
| NIHSS score at baseline | 1.124(1.032-1.224) | 0.007* | ... | ... |
| Dysphagia | 2.301(1.230-4.306) | 0.009* | ... | ... |
| A2DS2 score | ... | ... | 1.439(1.250-1.656) | <0.001*** |
| Thrombolysis |  | 0.674 |  | 0.468 |
| Leukocyte count | 1.194(1.047-1.360) | 0.008* | 1.165(1.021-1.330) | 0.023* |
| NLR |  | 0.070 |  | 0.070 |
| Hs-CRP | 1.015(1.005-1.026) | 0.005* | 1.014(1.004-1.025) | 0.007* |
| Albumin |  | 0.659 |  | 0.698 |
| FPR >18.22 | 3.028(1.607-5.706) | 0.001* | 3.239(1.727-6.075) | <0.001*** |

Abbreviations: SAP, stroke-associated pneumonia; OR: odds ratio; CI: confidence interval; NIHSS, National Institutes of Health Stroke Scale; The A2DS2 score includes age (1 point for $\geq 75$ years), atrial fibrillation (1 point), dysphagia (2 points), male sex (1 point), and stroke severity (0 point for NIHSS score 0-4, 3 points for NIHSS score 5-15, and 5 points for NIHSS score $\geq 16$ ); NLR, neutrophil to lymphocyte ratio; Hs-CRP, high-sensitivity C-reactive protein; FPR, fibrinogen-prealbumin ratio
†Adjusted with $P < 0.05$ in univariate analysis
‡Adjusted with $P < 0.05$ in the univariate analysis and A2DS2
\*p values considered statistically significant; \* $p < 0.05$ , \*\*\* $p < 0.001$

**Supplemental Table II.**
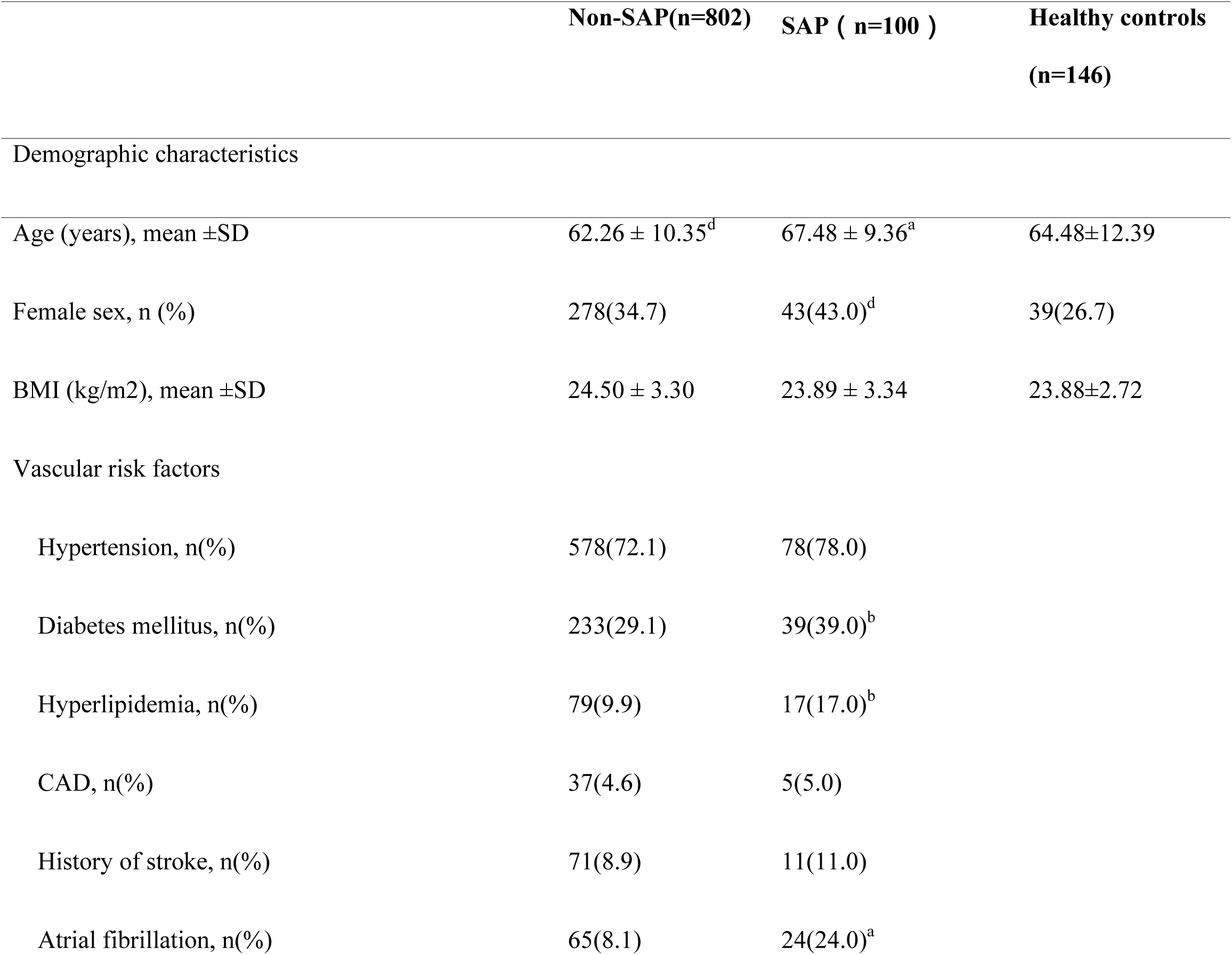
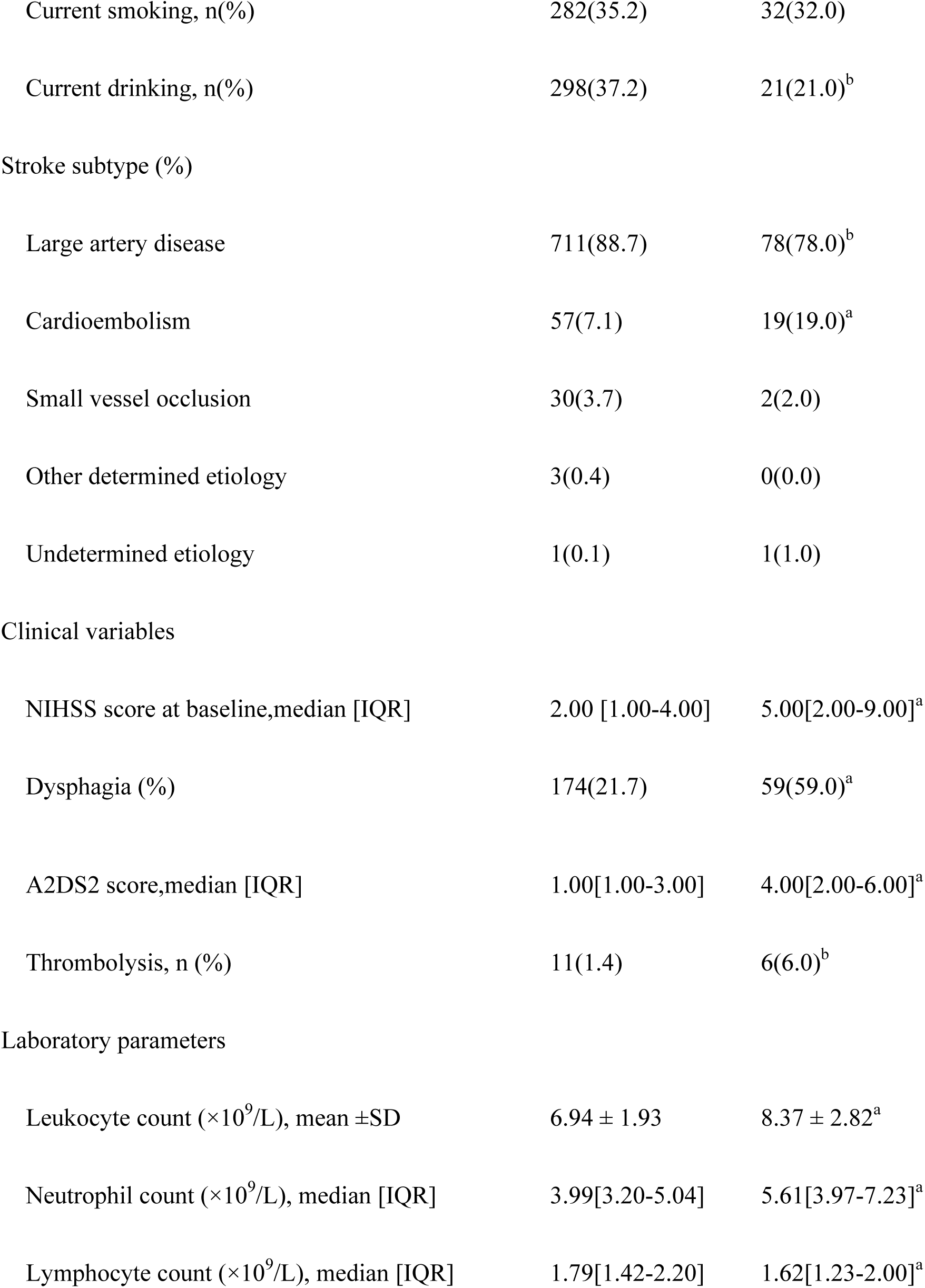
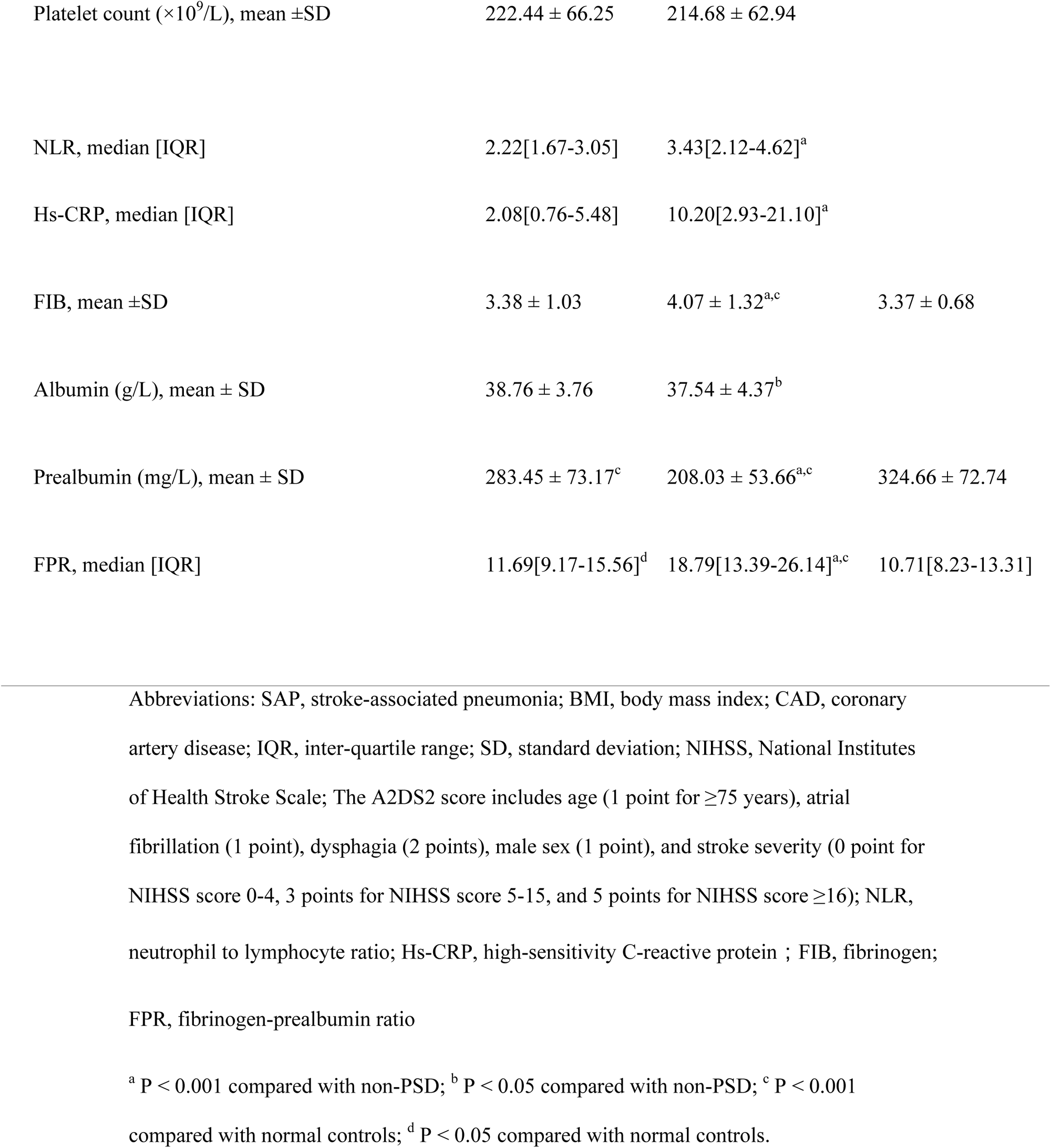
Clinical and demographic characteristics of the samples under study, defining only definite SAP as an outcome variable

|  | Non-SAP(n=802) | SAP ( n=100 ) | Healthy controls<br>(n=146) |
| --- | --- | --- | --- |
| Demographic characteristics |  |  |  |
| Age (years), mean ±SD | 62.26 ± 10.35<sup>d</sup> | 67.48 ± 9.36<sup>a</sup> | 64.48±12.39 |
| Female sex, n (%) | 278(34.7) | 43(43.0)<sup>d</sup> | 39(26.7) |
| BMI (kg/m<sup>2</sup>), mean ±SD | 24.50 ± 3.30 | 23.89 ± 3.34 | 23.88±2.72 |
| Vascular risk factors |  |  |  |
| Hypertension, n(%) | 578(72.1) | 78(78.0) |  |
| Diabetes mellitus, n(%) | 233(29.1) | 39(39.0)<sup>b</sup> |  |
| Hyperlipidemia, n(%) | 79(9.9) | 17(17.0)<sup>b</sup> |  |
| CAD, n(%) | 37(4.6) | 5(5.0) |  |
| History of stroke, n(%) | 71(8.9) | 11(11.0) |  |
| Atrial fibrillation, n(%) | 65(8.1) | 24(24.0)<sup>a</sup> |  |

|  |  |  |
| --- | --- | --- |
| Current smoking, n(%) | 282(35.2) | 32(32.0) |
| Current drinking, n(%) | 298(37.2) | 21(21.0)<sup>b</sup> |
| Stroke subtype (%) |  |  |
| Large artery disease | 711(88.7) | 78(78.0)<sup>b</sup> |
| Cardioembolism | 57(7.1) | 19(19.0)<sup>a</sup> |
| Small vessel occlusion | 30(3.7) | 2(2.0) |
| Other determined etiology | 3(0.4) | 0(0.0) |
| Undetermined etiology | 1(0.1) | 1(1.0) |
| Clinical variables |  |  |
| NIHSS score at baseline, median [IQR] | 2.00 [1.00-4.00] | 5.00[2.00-9.00]<sup>a</sup> |
| Dysphagia (%) | 174(21.7) | 59(59.0)<sup>a</sup> |
| A2DS2 score, median [IQR] | 1.00[1.00-3.00] | 4.00[2.00-6.00]<sup>a</sup> |
| Thrombolysis, n (%) | 11(1.4) | 6(6.0)<sup>b</sup> |
| Laboratory parameters |  |  |
| Leukocyte count ($\times 10^9/L$), mean ±SD | 6.94 ± 1.93 | 8.37 ± 2.82<sup>a</sup> |
| Neutrophil count ($\times 10^9/L$), median [IQR] | 3.99[3.20-5.04] | 5.61[3.97-7.23]<sup>a</sup> |
| Lymphocyte count ($\times 10^9/L$), median [IQR] | 1.79[1.42-2.20] | 1.62[1.23-2.00]<sup>a</sup> |
| Platelet count ($\times 10^9/L$), mean ±SD | 222.44 ± 66.25 | 214.68 ± 62.94 | |
| NLR, median [IQR] | 2.22[1.67-3.05] | 3.43[2.12-4.62]<sup>a</sup> |  |
| Hs-CRP, median [IQR] | 2.08[0.76-5.48] | 10.20[2.93-21.10]<sup>a</sup> |  |
| FIB, mean ±SD | 3.38 ± 1.03 | 4.07 ± 1.32<sup>a,c</sup> | 3.37 ± 0.68 |
| Albumin (g/L), mean ± SD | 38.76 ± 3.76 | 37.54 ± 4.37<sup>b</sup> | |
| Prealbumin (mg/L), mean ± SD | 283.45 ± 73.17<sup>c</sup> | 208.03 ± 53.66<sup>a,c</sup> | 324.66 ± 72.74 |
| FPR, median [IQR] | 11.69[9.17-15.56]<sup>d</sup> | 18.79[13.39-26.14]<sup>a,c</sup> | 10.71[8.23-13.31] |

Abbreviations: SAP, stroke-associated pneumonia; BMI, body mass index; CAD, coronary artery disease; IQR, inter-quartile range; SD, standard deviation; NIHSS, National Institutes of Health Stroke Scale; The A2DS2 score includes age (1 point for $\geq 75$ years), atrial fibrillation (1 point), dysphagia (2 points), male sex (1 point), and stroke severity (0 point for NIHSS score 0-4, 3 points for NIHSS score 5-15, and 5 points for NIHSS score $\geq 16$ ); NLR, neutrophil to lymphocyte ratio; Hs-CRP, high-sensitivity C-reactive protein ; FIB, fibrinogen; FPR, fibrinogen-prealbumin ratio
<sup>a</sup> $P < 0.001$ compared with non-PSD; <sup>b</sup> $P < 0.05$ compared with non-PSD; <sup>c</sup> $P < 0.001$ compared with normal controls; <sup>d</sup> $P < 0.05$ compared with normal controls.

**Supplemental Table III.**
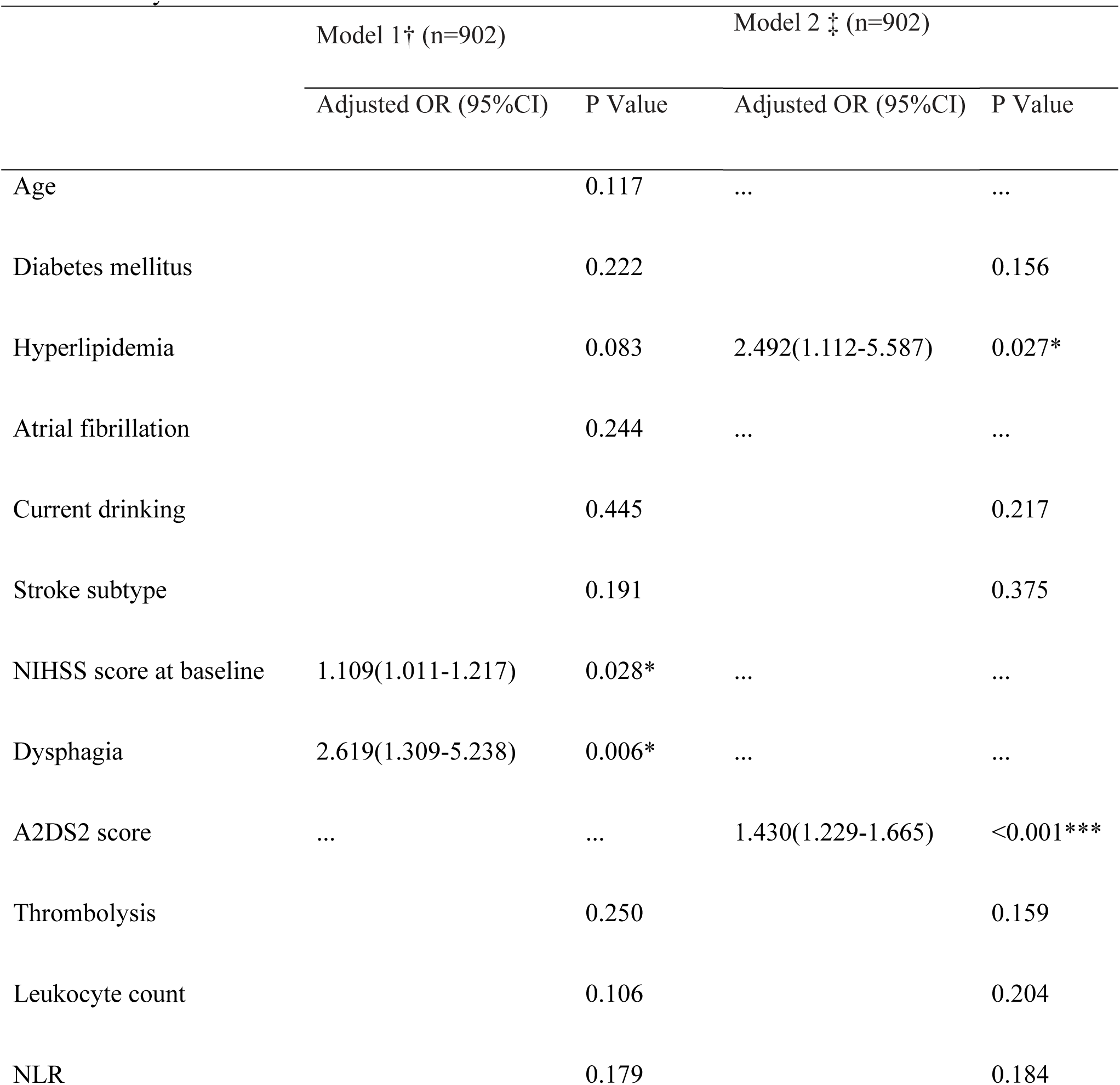
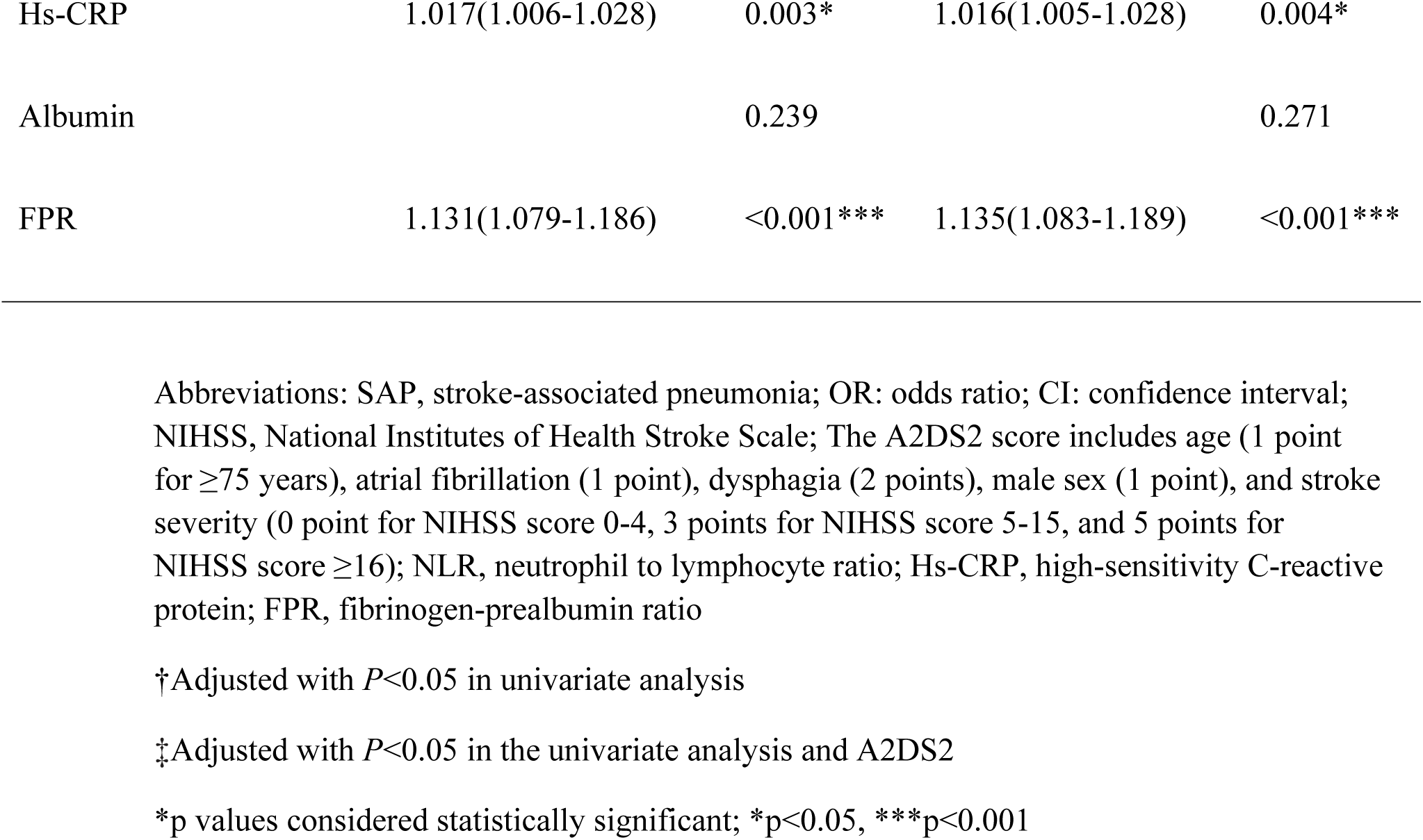
Binary logistic model of the clinical determinants of SAP, defining only definite SAP as an outcome variable

## Notes

### Competing Interest Statement

The authors have declared no competing interest.

